# Estimating the impact of physical distancing measures in containing COVID-19: an empirical analysis

**DOI:** 10.1101/2020.06.11.20128074

**Authors:** Wee Chian Koh, Lin Naing, Justin Wong

**Author notes:** Corresponding author: Dr Wee Chian Koh, Centre for Strategic and Policy Studies, Simpang 347, Jalan Gadong, Bandar Seri Begawan BE1318, Brunei Darussalam. **Funding** This research did not receive any specific grant from funding agencies in the public, commercial, or not-for-profit sectors. **Ethical approval** Not required. **Authors’ contributions** WCK conceived and designed the study, collected the data, and conducted the statistical analyses. WCK and JW wrote the manuscript with critical feedback from LN. All authors contributed to and approved the final manuscript.

## Abstract

**Background:** Epidemic modelling studies predict that physical distancing is critical in containing COVID-19. However, few empirical studies have validated this finding. Our study evaluates the effectiveness of different physical distancing measures in controlling viral transmission.

**Methods:** We identified three distinct physical distancing measures with varying intensity and implemented at different times—international travel controls, restrictions on mass gatherings, and lockdown-type measures—based on the Oxford COVID-19 Government Response Tracker. We also estimated the time-varying reproduction number (R_t_) for 142 countries and tracked R_t_ temporally for two weeks following the 100^th^ reported case in each country. We regressed R_t_ on the physical distancing measures and other control variables (income, population density, age structure, and temperature) and performed several robustness checks to validate our findings.

**Findings:** Complete travel bans and all forms of lockdown-type measures have been effective in reducing average R_t_ over the 14 days following the 100^th^ case. Recommended stay-at-home advisories and partial lockdowns are as effective as complete lockdowns in outbreak control. However, these measures have to be implemented early to be effective. Lockdown-type measures should be instituted two weeks before the 100th case and travel bans about a week before detection of the first case.

**Interpretation:** A combination of physical distancing measures, if implemented early, can be effective in containing COVID-19—tight border controls to limit importation of cases, encouraging physical distancing, moderately stringent measures such as working from home, and a full lockdown in the case of a probable uncontrolled outbreak.

**Research in context:** *Evidence before this study:* Evidence on the impact of physical distancing measures on containing COVID-19 has primarily relied on epidemic modelling studies. As cases accumulate worldwide, it has become possible to use empirical data to validate the model-based findings. The few empirical studies that analyze global case data find that lockdowns and international travel restrictions are important, but have not explored, beyond these broad findings, the intensity and timeliness of the various measures to inform policymaking.

*Added value of this study:* We assessed, at a normalized stage of the epidemic curve, how the intensity and implementation timing of various physical distancing measures in 142 countries affect viral transmission, measured by the time-varying reproduction number (R_t_). Other similar empirical studies treat the measures as binary variables, do not address the potential confounding effect of increased caseload on transmission, and do not use R_t_ as the primary metric.

*Implications of all the available evidence:* Our results support the findings in modelling studies, and subsequent empirical studies, that physical distancing measures can limit disease spread. We found that full border control and complete lockdowns are effective, but less stringent measures such as stay-at-home recommendations and working from home are as effective. As such, the framing of lockdown measures as a binary approach may be counterproductive. Overall, these measures are only effective if they are implemented early.

## Introduction

Coronavirus disease 2019 (COVID-19) is an emerging respiratory infectious disease caused by the severe acute respiratory syndrome coronavirus 2 (SARS-CoV-2), which was first detected in early December 2019 in Wuhan, China. As of May 31, 2020, it has affected 5.93 million people and resulted in more than 367,000 deaths globally (WHO 2020).

In the absence of effective therapeutics or vaccines, containment measures rely on the capacity to control viral transmission through non-pharmaceutical interventions (NPIs) (Kissler et al. 2020). Current evidence suggests that the effectiveness of case isolation and contact tracing strategies can be enhanced when combined with physical distancing measures in public settings (Chu et al. 2020; Kucharski et al. 2020). Governments worldwide have implemented various forms of physical distancing measure with varied stringency level and timeliness. The measures include school and workplace closures, cancellation of public events, restrictions on mass gatherings, public transport closures, stay-at-home orders, restrictions on internal movements, and international travel controls. Due to the potential for socioeconomic disruptions caused by these measures, it is therefore important to quantify their impact on disease spread to inform policymaking, which has thus far relied primarily on epidemic modelling studies (Ferguson et al. 2020; Prem et al. 2020). As cases accumulate, it has become possible to use empirical data derived from real-world observations to validate the model-based estimates of the effectiveness of policy interventions.

In this paper, we assessed—at a normalized point on the epidemic curve—the impact of physical distancing measures on viral transmission measured by the time-varying reproduction number, R_t_, which represents the expected number of secondary cases generated by a primary case at time t. A value of R_t_ greater than one indicates that a sustained outbreak is likely. The goal of policy intervention is to bring R_t_ below one, suggesting that the outbreak is under control.

## Methods

### Physical distancing measures

Data on physical distancing measures were obtained from the Oxford COVID-19 Government Response Tracker (OxCGRT), which collects information, starting from January 1, 2020, on a range of government policies, assigns a stringency score for the measures, and aggregates the data into a common index for 170 countries (May 28 version). We used the Stringency Index as an aggregate measure, which has a score between 0 and 100, with a higher index indicating increased stringency. We further examined the impact of specific measures: (i) school closures; (ii) workplace closures; (iii) cancellation of public events; (iv) restrictions on size of gatherings; (v) public transport closures; (vi) stay-at-home orders; (vii) restrictions on internal movements, and (viii) restrictions on international travel. These measures have an ordinal scale of severity or intensity. Further details on the OxCGRT database are provided in Hale et al. (2020).

### Estimation of the real-time reproduction number

We normalized the stage of disease spread to minimize the confounding effect of increased caseload on transmission: the impact of interventions is expected to be different at 10 and 1,000 cases. We used 100 total cases as the starting point for all countries to indicate an outbreak (Hartfield and Alizon 2013).

We estimated R_t_ for 142 countries that have reported at least 100 cases as of May 28, 2020. The estimation covered the whole period from the first reported case to May 28 using a weekly sliding window based on the methods developed by Cori et al. (2013). We used data on new daily cases and the distribution of the generation time (time between infection of an index case and infection of a secondary case). We incorporated uncertainty in the generation time distribution with a mean of 3.6 days (sd: 0.7 days) and standard deviation of 3 days (sd: 0.8 days) and used a Gamma prior for the reproduction number with mean 2.6 and standard deviation 2. These parameter estimates were obtained from the COVID-19 Epiforecasts project by the Centre for the Mathematical Modelling of Infectious Diseases at the London School of Hygiene and Tropical Medicine (see Abbott et al. 2020). Data on daily reported cases were obtained from the European Centre for Disease Prevention and Control and from the Johns Hopkins University Centre for Systems Science and Engineering COVID-19 Data Depository. R_t_ was estimated using the EpiEstim package in R version 3.6.3 (R Foundation for Statistical Computing).

An important feature of examining R_t_, instead of cumulative case numbers, is that, if the proportion of cases that are unreported remain constant throughout an outbreak, estimates of R_t_ are unaffected by underreporting (Thompson et al. 2019).

### Regression model

As countries have implemented and subsequently relaxed measures in response to the outbreak, establishing causality from such measures to a change in R_t_ is challenging. To address possible reverse causality, we examined the measures that were in place at the time when 100 cases have been reported. We then tracked R_t_ temporally over the next 14 days. The lagged measures thus controls for the endogenous response to viral transmission.

We regressed R_t_ on physical distancing measures and other covariates. The control variables used were income level (log of GDP per capita at current US$), population density (log of population per square kilometre), age structure (proportion of population aged 65 years and above), and air temperature (14-day average after the 100^th^ case). These socioeconomic and environmental factors have been postulated to influence disease spread (Liu et al. 2020; Qiu et al. 2020). Data on GDP per capita, population density, and population above 65 years old were obtained from the World Bank’s World Development Indicators, supplemented by the Central Intelligence Agency’s The World Factbook. Data on temperature were collected from the Air Quality Open Data Platform and other online weather resources.

The empirical specification takes the following form:

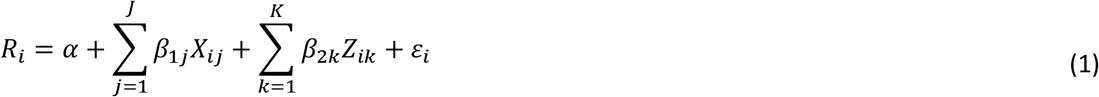

Where 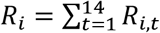 is the average reproduction number of country over the 14 days following the date of the 100^th^ case; *X*_*ij*_ is country *l*’s physical distancing measure of type *j* on the date of the 100^th^ case;*Z*_*ik*_ represents the country characteristic *k* (income level, population density, age structure, and temperature) of country; *α* is a constant term, *β*’s are the regression coefficients, and *ε*_*i*_ denotes the error term.

We also conducted ex-post predictions on the date of the 100^th^ case using equation (1) to make comparative assessments on how R_t_ would be predicted to evolve relative to what has been observed in reality. All regression analyses were performed in Stata 14 (StataCorp LLC).

### Robustness checks

We conducted a series of robustness checks to validate our results. We explored a shorter time horizon of seven days to address the possibility of new measures implemented after the 100^th^ case that could affect R_t._ We also used the growth in total cases instead of R_t_ as the dependent variable. To examine actual behavioural changes instead of de jure government policies, we used a de facto measure of physical distancing using mobility data from Google Community Mobility Reports.

## Results

We first take a cursory look at the nature of the relationship between physical distancing measures and R_t_, and then proceed to estimate the magnitudes using regression models.

Figure 1 shows how R_t_ (average over the 14 days following the 100^th^ case) varies with the stringency of physical distancing measures (as of date of 100^th^ case). There are several important observations: (i) countries with more stringent measures tend to have lower R_t_ on average, as illustrated by the downward sloping line; (ii) no country with a stringency index lower than 50 could bring average R_t_ to below one within two weeks; and (iii) countries with R_t_ less than 1.5 on the date of the 100^th^ case have generally kept total cases (size of the bubbles) at a manageable level (as of May 28).

**Figure 1.**
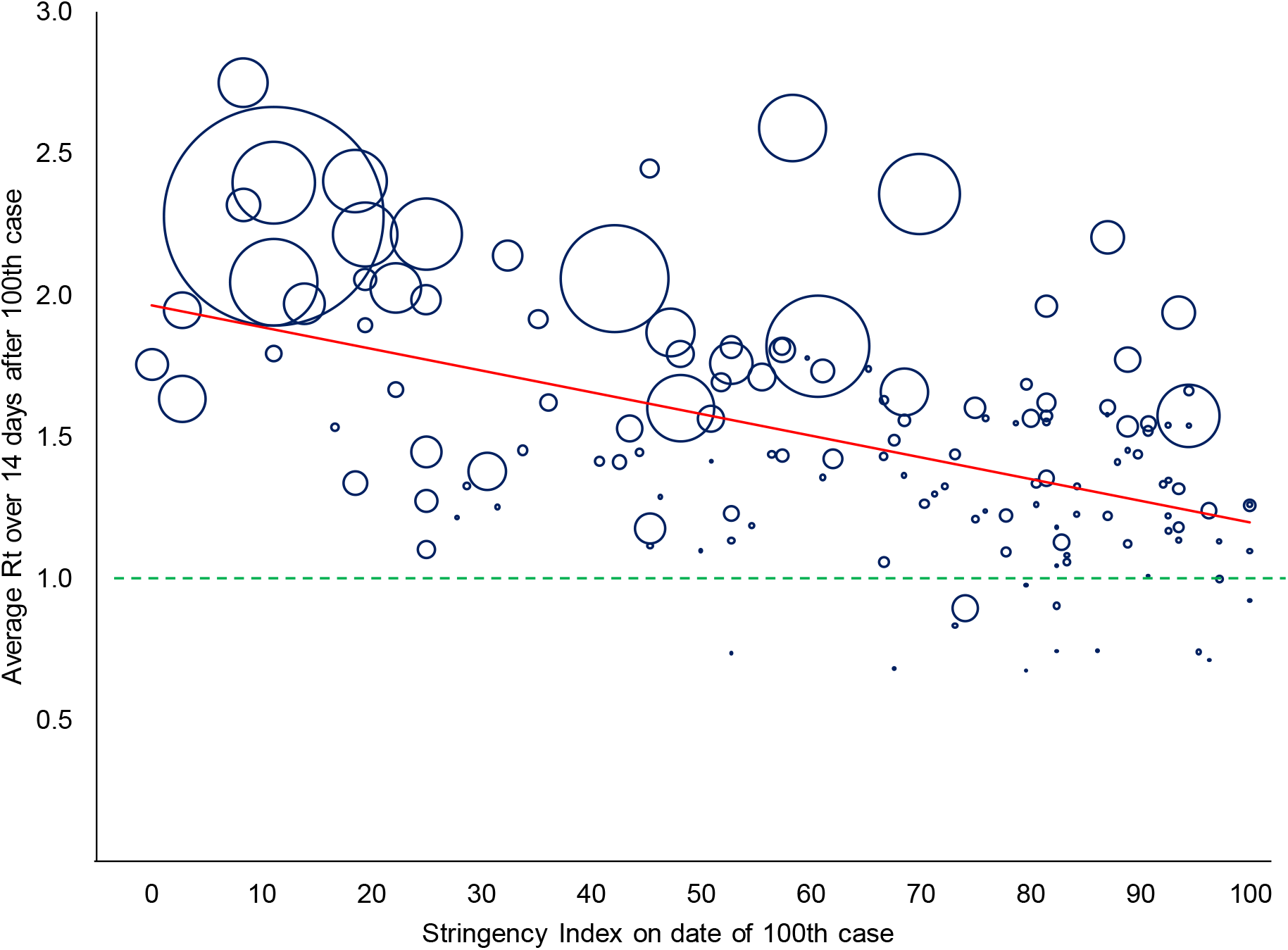
Stringency index of physical distancing measures on the date of the 100^th^ case and average reproduction numbers in the following two weeks. The time-varying reproduction number R_t_ is the expected number of secondary cases generated by a primary case at time t. The Stringency Index is a composite index of physical distancing measures with a range of 0 to 100, calculated by the Oxford COVID-19 Government Response Tracker (OxCGRT); a larger value indicates higher stringency. Each bubble represents a country, and the size of the bubble is proportional to the total number of reported cases as of May 28, 2020. The red solid line is the best linear fit of the relationship between the stringency level on the date of the 100^th^ reported case and the average R t in the following two weeks. The green dashed line is the R_t_ threshold: a value below one indicates that a sustained outbreak is unlikely if the measures remain in place.

The average timing of implementation of the physical distancing measures is summarized in Table S1 in the Supplementary materials. The earliest policies to be implemented, on average, were restrictions on international travel, about 11 days before the detection of the first case. Cancellation of public events and school closures were the initial responses during the onset of an outbreak (about a week after the first case), followed by restrictions on the size of gatherings and more stringent measures such as workplace closures, restrictions on internal movement, stay-at-home orders, and public transport closures. On average, all these measures were implemented before the occurrence of the 100^th^ case.

Since several measures were implemented very close to one another, and due to the similar nature of some the measures, it is difficult to relate the observed changes in R_t_ to a specific measure. We addressed this identification issue by grouping the measures, taking into account the implementation timing and correlation (Table S2 in Supplementary materials). We identified three distinct categories of physical distancing measures amenable to our analysis: (i) restrictions on international travel; (ii) restrictions on mass gatherings; and (iii) lockdown-type measures. Within each category, the intensity and timeliness of implementation varies (Table 1). In particular, we highlight the wide range of measures within a category. For instance, in lockdown-type measures, the least stringent being recommendations and government advisories on internal movement, up to complete lockdown with closure of all non-essential workplaces and stay-at-home requirements with minimal exceptions.

**Table 1.**
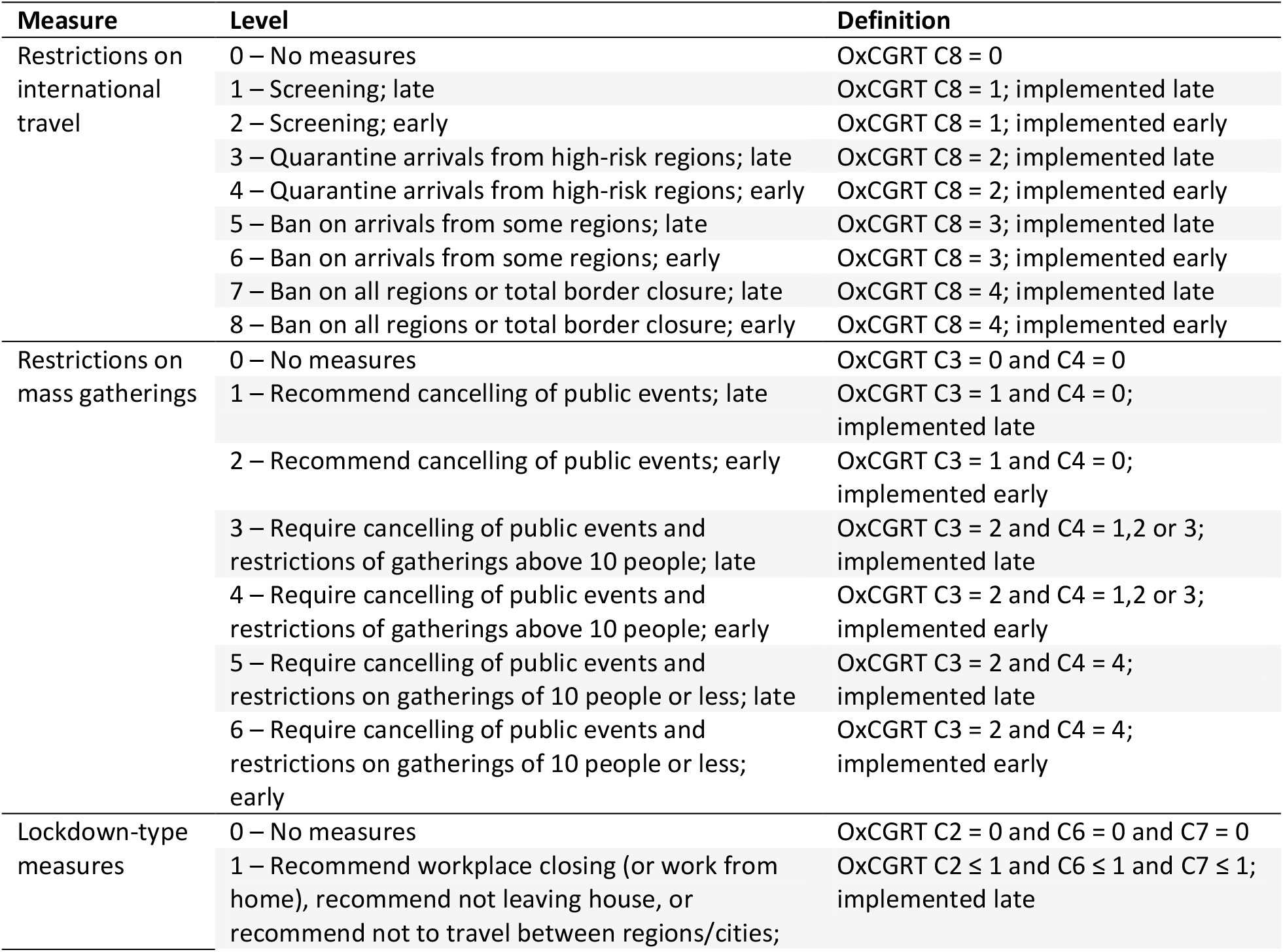
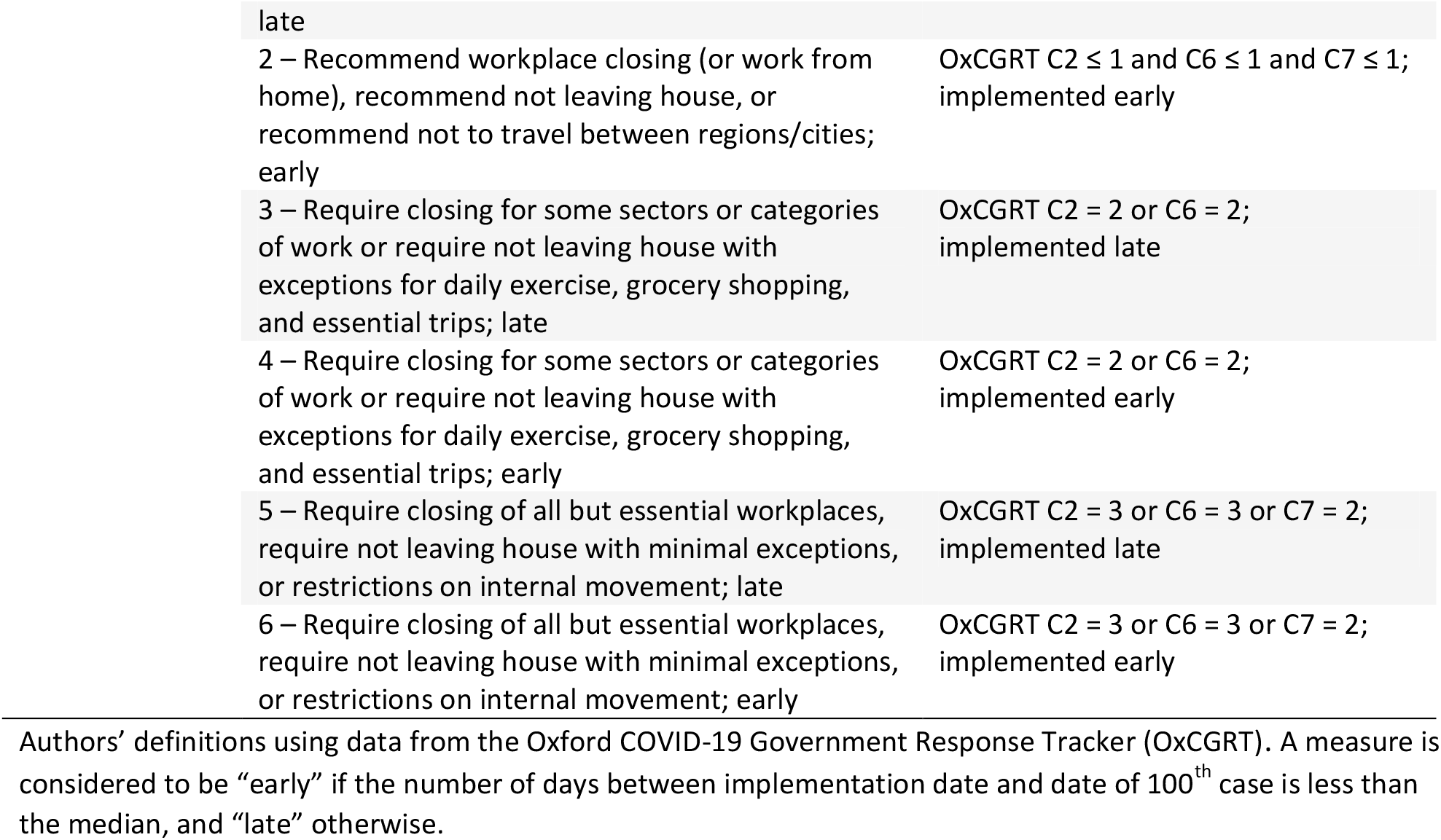
Physical distancing measures.

Restrictions on international travel are based on OxCGRT indicator C8. Restrictions on mass gatherings combine OxCGRT indicators C3 (cancel public events) and C4 (size restriction of gatherings). Lockdown combines OxCGRT indicators C2 (workplace closures), C6 (stay-at-home requirements), and C7 (restrictions on internal movement). A measure is considered to be implemented early if the number of days between implementation date and date of 100^th^ case is less than the global median, and late otherwise. For instance, Taiwan acted swiftly in banning arrivals for some regions (56 days before 100^th^ reported case, compared to global median of 31 days before 100^th^ case) while Sweden was late to implement border controls (12 days after 100^th^ case).

Figures 2 to 4 show how R_t_ varies with each physical distancing measure. In general, the more stringent the measure and the earlier its implementation, the lower the value of R_t_. In the case of international travel restrictions, R_t_ below one was only observed in countries that either implemented quarantine of arriving passengers from high-risk regions early, or enacted bans on arrivals. On mass gathering restrictions, R_t_ less than one was observed in countries that either cancelled public events or limited the size of gatherings. Interestingly, one country (Brunei) managed to bring R_t_ to below one without the need for any form of lockdown-type measures. Overall, countries with earlier and more stringent measures at the time of the 100^th^ case appeared to have recorded far fewer cases (as of May 28), although there are a few exceptions (e.g. Peru and Russia, despite early lockdowns).

**Figure 2.**
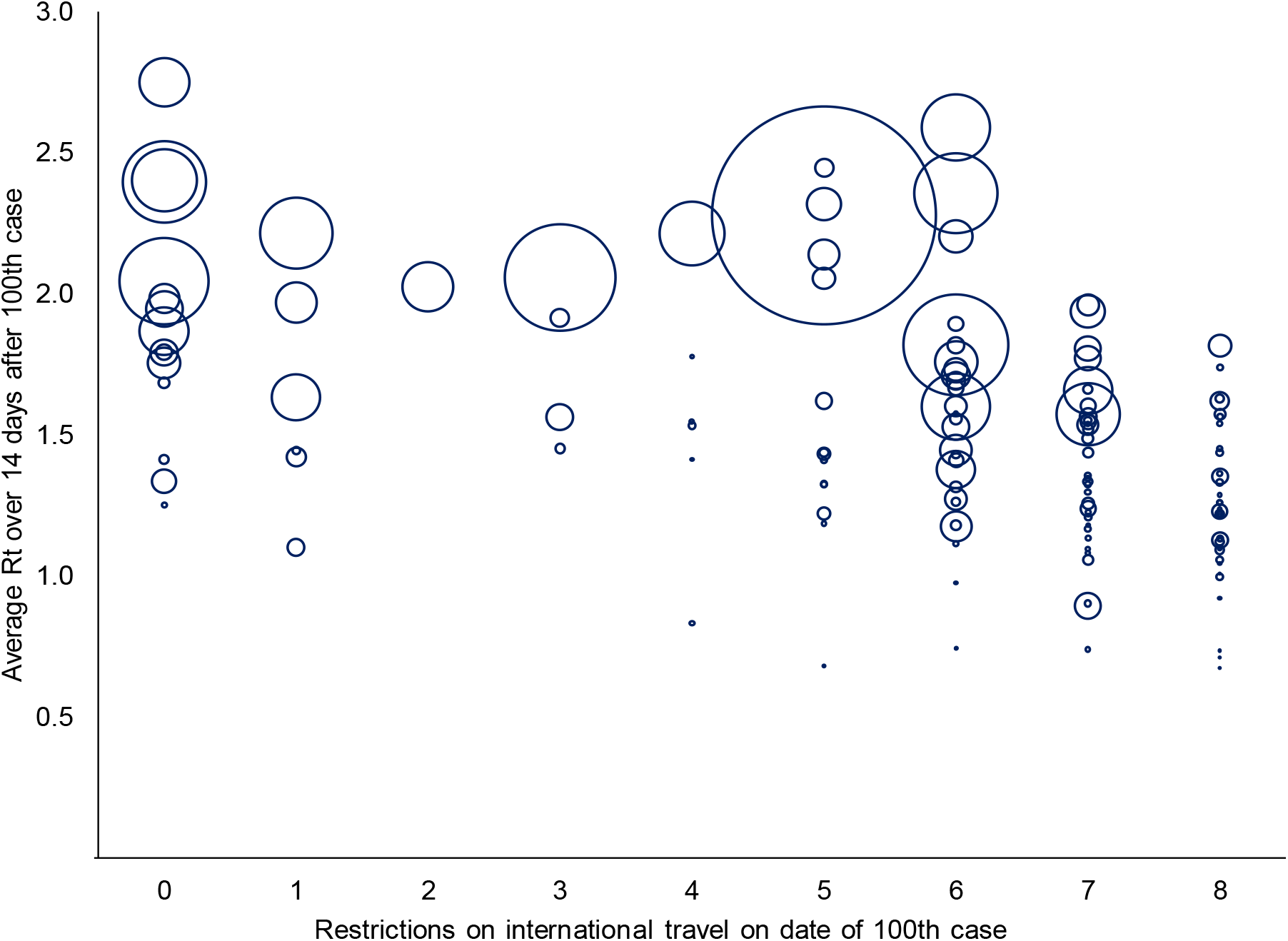
Restrictions on international travel on the date of the 100^th^ case and average reproduction numbers in the following two weeks. The time-varying reproduction number R_t_ is the expected number of secondary cases generated by a primary case at time t. The specific measures on international travel restrictions are detailed in Table 1. Each bubble represents a country, and the size of the bubble is proportional to the total number of reported cases as of May 28, 2020.

**Figure 3.**
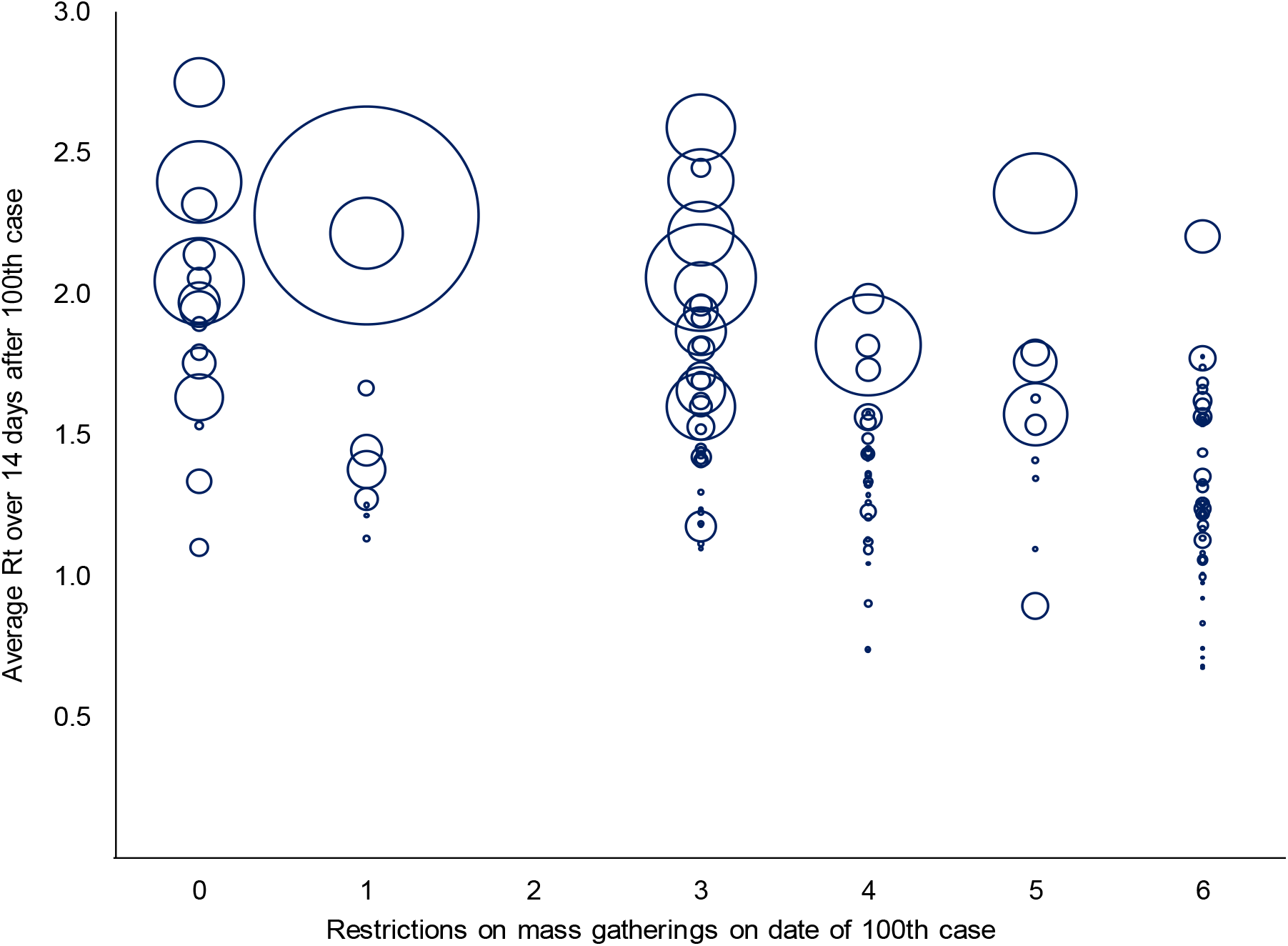
Restrictions on mass gatherings on the date of the 100^th^ case and average reproduction numbers in the following two weeks. The time-varying reproduction number R_t_ is the expected number of secondary cases generated by a primary case at time t. The specific measures on mass gathering restrictions are detailed in Table 1. Each bubble represents a country, and the size of the bubble is proportional to the total number of reported cases as of May 28, 2020.

**Figure 4.**
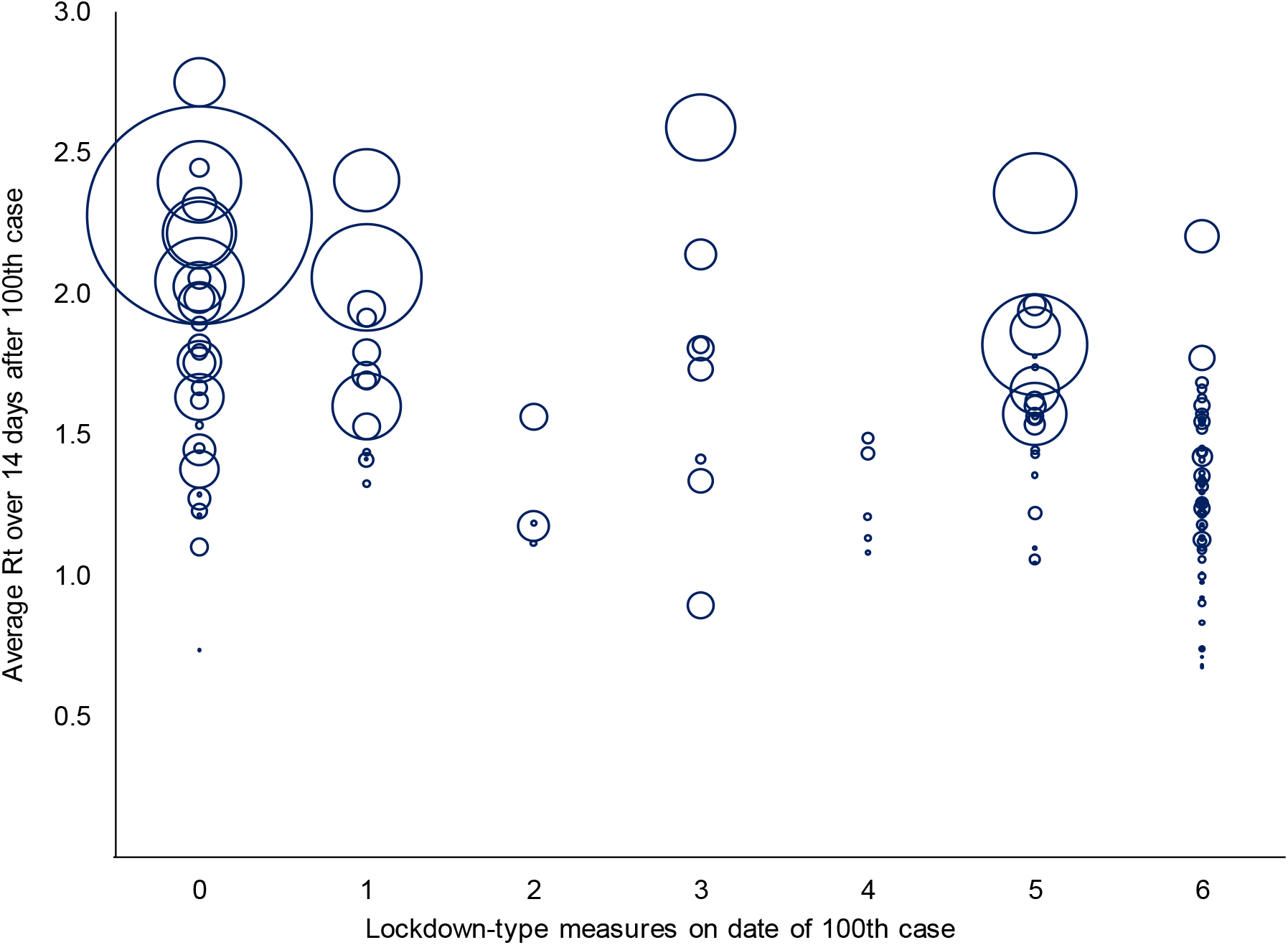
Lockdown-type measures on the date of the 100^th^ case and average reproduction numbers in the following two weeks. The time-varying reproduction number R_t_ is the expected number of secondary cases generated by a primary case at time t. The specific measures on lockdown are detailed in Table 1. Each bubble represents a country, and the size of the bubble is proportional to the total number of reported cases as of May 28, 2020.

### Impact of physical distancing measures

We first examine the impact of the Stringency Index on R_t_. The results are reported in column (1) of Table 2. The estimated coefficient of the Stringency Index is negative and significant: an increase in the index by 10 points reduces R_t_ by 0.06 (95% CI: −0.08, −0.04). Among the other independent variables, warmer temperature is associated with a lower R_t_: an increase in the temperature of 10 degree Celsius reduces R_t_ by 0.16 (95% CI: −0.24, −0.09).

**Table 2.** Estimated impact of the Stringency Index on COVID-19 transmission.

|  | Dependent variable |  |  |  |
| --- | --- | --- | --- | --- |
| | $R_{t,14}$ | $R_{t,7}$ | $g$ | $R_{t,14}$ |
| Stringency Index | -0.0061***<br>(0.0012) | -0.0072***<br>(0.0017) | -0.1946***<br>(0.0454) |  |
| Google mobility |  |  |  | 0.0052***<br>(0.0015) |
| ln GDP per capita | -0.0412<br>(0.0285) | -0.0427<br>(0.0399) | -0.2865<br>(1.0912) | -0.0053<br>(0.0325) |
| ln population density | -0.0126<br>(0.0213) | -0.0181<br>(0.0299) | 0.1416<br>(0.8179) | -0.0148<br>(0.0245) |
| % age 65 in population | 0.0006<br>(0.0072) | 0.0018<br>(0.0101) | -0.0118<br>(0.2776) | 0.0034<br>(0.0083) |
| Temperature | -0.0162***<br>(0.0039) | -0.0184***<br>(0.0054) | -0.4665***<br>(0.1478) | -0.0136***<br>(0.0050) |
| Constant | 2.5674***<br>(0.2700) | 2.8634***<br>(0.3780) | 32.4252***<br>(10.3474) | 1.9881***<br>(0.3084) |
| Number of countries | 142 | 142 | 142 | 102 |
| $R^2$ | 0.388 | 0.314 | 0.314 | 0.353 |
| Adjusted $R^2$ | 0.366 | 0.289 | 0.289 | 0.320 |

The dependent variable in columns (1) and (4), $R_{t,14}$ is the average $R_t$ over the 14 days following the date of the 100<sup>th</sup> case; $R_{t,7}$ in column (2) is the average $R_t$ over the 7 days following the date of the 100<sup>th</sup> case; $g$ in column (3) is the growth rate of total cases between the date of the 100<sup>th</sup> case and the date 14 days later. The Stringency index is a composite index of physical distancing measures with a range of 0 to 100, calculated by the Oxford COVID-19 Government Response Tracker (OxCGRT). Mobility (from Google Community Mobility Reports) is the average percent change in visits to retail and recreation, grocery and pharmacy, parks, transit and workplaces on the date of the 100<sup>th</sup> case compared to the median baseline value of the corresponding day of the week during January 3 to February 6, 2020, as a proxy measurement of de facto physical distancing; a positive coefficient indicates that a reduction in mobility reduces $R_{t,14}$ .

We then assess the impact of the three categories of physical distancing measures individually, and then collectively. Table 3 reports the results. Columns (1) to (3) affirm the observations from Figures 2 to 4 that earlier and more stringent measures were associated with lower R_t_. However, some of the significant results disappeared when all the variables were entered simultaneously.

**Table 3.** Estimated impact of the type of physical distancing measures on COVID-19 transmission.

|  | (1) | (2) | (3) | (4) |
| --- | --- | --- | --- | --- |
| Restrictions on international travel |  |  |  |  |
| 1 | -0.1709<br>(0.1688) |  |  | -0.2678<br>(0.1602) |
| 2 | -0.2877<br>(0.3683) |  |  | -0.3115<br>(0.3604) |
| 3 | 0.1086<br>(0.2027) |  |  | 0.1362<br>(0.2041) |
| 4 | -0.1742<br>(0.1741) |  |  | -0.1264<br>(0.1682) |
| 5 | -0.1339<br>(0.1337) |  |  | -0.0530<br>(0.1302) |
| 6 | -0.1433<br>(0.1201) |  |  | -0.0763<br>(0.1214) |
| 7 | -0.3188***<br>(0.1189) |  |  | -0.1391<br>(0.1256) |
| 8 | -0.4133***<br>(0.1242) |  |  | -0.2432*<br>(0.1288) |
| Restrictions on mass gatherings |  |  |  |  |
| 1 |  | -0.2361<br>(0.1435) |  | -0.1620<br>(0.1482) |
| 2 |  | N/A |  | N/A |
| 3 |  | -0.1495<br>(0.1059) |  | -0.0308<br>(0.1313) |
| 4 |  | -0.4142***<br>(0.1162) |  | -0.2064<br>(0.1379) |
| 5 |  | -0.2746**<br>(0.1366) |  | -0.1094<br>(0.1595) |
| 6 |  | -0.4791***<br>(0.1073) |  | -0.2124<br>(0.1471) |
| Lockdown-type measures |  |  |  |  |
| 1 |  |  | -0.0368<br>(0.1094) | -0.1366<br>(0.1264) |
| 2 |  |  | -0.3318*<br>(0.1747) | -0.4465**<br>(0.1879) |
| 3 |  |  | -0.1107<br>(0.1297) | -0.1847<br>(0.1391) |
| 4 |  |  | -0.5125***<br>(0.1574) | -0.3774**<br>(0.1714) |
| 5 |  |  | -0.1586<br>(0.0954) | -0.1083<br>(0.1212) |
| 6 |  |  | -0.4268***<br>(0.0838) | -0.3186***<br>(0.1154) |
| In GDP per capita | -0.0207<br>(0.0291) | -0.0389<br>(0.0294) | -0.0505*<br>(0.0290) | -0.0642**<br>(0.0307) |
| In population density | -0.0191<br>(0.0231) | -0.0139<br>(0.0221) | -0.0059<br>(0.0213) | -0.0063<br>(0.0226) |
| % age 65 in population | -0.0026<br>(0.0077) | 0.0015<br>(0.0073) | 0.0047<br>(0.0072) | 0.0026<br>(0.0073) |
| Temperature | -0.0181***<br>(0.0044) | -0.0171***<br>(0.0039) | -0.0157***<br>(0.0038) | -0.0152***<br>(0.0041) |
| Constant | 2.3303***<br>(0.2766) | 2.4854***<br>(0.2819) | 2.4326***<br>(0.2687) | 2.7897***<br>(0.3117) |
| Number of countries | 142 | 142 | 142 | 142 |
| R<sup>2</sup> | 0.372 | 0.402 | 0.435 | 0.512 |
| Adjusted R<sup>2</sup> | 0.313 | 0.361 | 0.392 | 0.417 |

Column (4) shows that travel bans on all regions and lockdown-type measures, if implemented early, significantly lowered R_t._ Relative to no measures being taken, a total border closure reduces R_t_ by 0.24 (95% CI: −0.50, 0.01); policies that recommend working from home or staying home reduce R_t_ by 0.45 (95% CI: −0.82, −0.07); a partial lockdown reduces R_t_ by 0.38 (95% CI: −0.72, −0.04); and a complete lockdown reduces R_t_ by 0.32 (95% CI: −0.55, −0.09).

To validate our results, we conducted several robustness checks. We tracked R_t_ over seven days and used the growth of total cases instead of R_t_ as the dependent variable. We also used a de facto measurement of physical distancing using Google mobility data—as opposed to de jure government announced measures—to examine the actual observed behaviour changes on R_t_. The findings are largely unchanged, as reported in columns (2) to (4) in Table 2 and the Supplementary materials (Tables S3 and S4, Figure S1).

### Ex-post predictions

We conducted ex-post predictions using the regression model in column (4) of Table 3 to retrospectively assess how R_t_ would be predicted to evolve over the following two weeks, given the case history and physical distancing measures implemented on the date of the 100^th^ case, along with specific country characteristics. The predicted R_t_ of the 142 countries are displayed in Table S5 in the Supplementary materials. Some countries had a lower R_t_ over the two weeks following the 100^th^ case than predicted by the model, such as Japan, Brunei, Iceland, and Vietnam. By contrast, others, such as Turkey, Italy, and the United States had a higher R_t_.

## Discussion

We assessed, at a standardized stage of the outbreak at 100 cases, the impact of physical distancing measures on COVID-19 transmission, measured by R_t_, and found that, on average, they have been effective in reducing R_t_—if the measures were sufficiently stringent and implemented early. By using R_t_ as the primary metric of transmission, instead of cumulative case counts in other similar studies (Castex et al. 2020; Deb et al. 2020), we accounted for potential confounding effects caused by under-testing and underreporting of cases.

Our study provides empirical support to findings from modelling studies that highlight the role of physical distancing measures in containing COVID-19. We identified three distinct measures, implemented at different times—restrictions on international travel prior to the first reported case, restrictions on mass gatherings during the onset of an outbreak, and lockdowns at later stages.

Our analysis suggests a hierarchy of physical distancing measures that are effective in outbreak control. We found that lockdown-type measures had the largest effect on limiting viral transmission, followed by complete travel bans. These measures have to be implemented early to be effective—based on our definition of early implementation using the observed median timing across countries worldwide, lockdown measures should be instituted two weeks before the 100^th^ case and travel bans about a week before detection of the first case.

This accords with the findings in other studies that severe travel restrictions have been critical in slowing down infections in China (Kucharski et al. 2020) and around the world (Keita 2020), and also corroborates studies showing that lockdowns limited disease spread in Wuhan (Fang et al. 2020), Italy and Spain (Tobias 2020), and California (Friedson et al. 2020).

Importantly, our findings suggest that lockdowns measures should not be viewed in a binary approach. There is a wide range of lockdown-type measures from less stringent forms such as working from home up to complete movement restrictions, and all were shown to be effective in suppressing viral transmission. If implemented early, work from home and stay at home recommendations reduce Rt by 0.45 (95% CI: −0.82, −0.07); a partial lockdown reduces R_t_ by 0.38 (95% CI: −0.72, −0.04); and a complete lockdown reduces R_t_ by 0.32 (95% CI: −0.55, −0.09). Across these three grades of lockdown-type measures, the 95% CI of their effect sizes overlap suggesting no significant difference in effectiveness across these measures. This finding is replicated even when assessed against other indicators of outbreak control, such as the increase in cumulative cases. As such, we suggest that early on in the outbreak, complete lockdowns may be unnecessary to control viral transmission, given the availability of other equally effective and more sustainable approaches. This is particularly important for the poorest countries. More than four-fifths of low- and lower-middle income countries have imposed complete lockdowns at the time of 100 reported cases (compared to three-fifths in upper-middle income and less than one-third in high-income countries), with potentially severe socioeconomic consequences, having already been hit by the slump in global economic activity, including sharp declines in remittances, tourism receipts, and commodity revenues (World Bank 2020).

Measures that recommend workplace closures or staying at home have been effective, implying that voluntary physical distancing has played an important role. In the United States, the decrease in mobility has been found to be largely voluntary, reflecting greater awareness of risk (Maloney and Taskin 2020). Japan has achieved success without the need for a complete lockdown. Clear public health messaging and voluntary practice of physical distancing shaped by cultural norms such as mask wearing, avoiding handshakes, and keeping silence when taking public transport and during events such as funerals, have been critical in limiting disease spread (Sposato 2020).

Overall, our analysis suggests that a combination of physical distancing measures may yield the most beneficial outcomes: international travel restrictions to limit imported cases from high-risk regions, encouraging voluntary social distancing, moderate forms of lockdown-type measures such as working from home and only leaving the house for necessary activities, and complete lockdowns in areas or provinces with more severe outbreaks. The implementation timeliness of these measures invariably depends on the country-specific context, including public acceptance and institutional capacity.

Countries that have been relatively successful share these common elements. Despite an international travel hub and its close proximity to Wuhan, early border control and the practice of personal protective behaviours, including the use of face masks, contributed to Hong Kong’s success in controlling viral transmission (Wong et al. 2020). Taiwan and Brunei responded quickly by instituting border control and reassured the public by delivering timely information on the epidemic (Wang et al. 2020; Wong et al. 2020). Targeted lockdown-type measures in Vietnam, coupled with mask wearing and consistent public health messaging, helped to contain disease spread (Duc Huynh 2020).

Our study has several limitations. First, although we controlled for several country characteristics, our model could suffer from omitted variable bias as behavioural variables, such as mask wearing, were unaccounted for due to lack of data. Second, beyond physical distancing measures, other NPIs such as early case isolation and aggressive contact tracing and quarantine are critical elements of a successful containment strategy (Ferguson et al. 2020), which we could not control for, again due to the lack of data. Third, although a significant amount of effort has been put into the construction of the OxCGRT database with a global coverage and a systematic classification of government policies, there could be some reporting errors or data quality issues. Moreover, our country-level analysis may miss the variation of policies implemented at the city/county/province level. Nonetheless, the database is the most comprehensive to date.

## Conclusion

Physical distancing measures have been applied in arguably every country that is fighting COVID-19. Although modelling studies have shown the importance of physical distancing in stemming disease spread, few empirical studies have validated this finding. We provide empirical support and quantified the impact of physical distancing measures in lowering the reproduction number, particularly lockdown-type measures and border closures. Moreover, we found that less stringent lockdown-type measures, such as encouraging working from home and staying home unless necessary were as effective as complete lockdowns in reducing transmission. However, all these measures have to be implemented early to be effective. As many countries are in the midst of de-escalating, we suggest that some combination of these measures—empirically justified—should be considered in containing subsequent waves of COVID-19.

## Data Availability

The datasets constructed for this paper can be made available upon request from the corresponding author.

## Acknowledgements

We thank Dr Ying-Ru Lo, Head of Mission and WHO Representative to Malaysia, Brunei Darussalam and Singapore, for useful comments and suggestions.

## Supplementary materials

**Figure S1.**
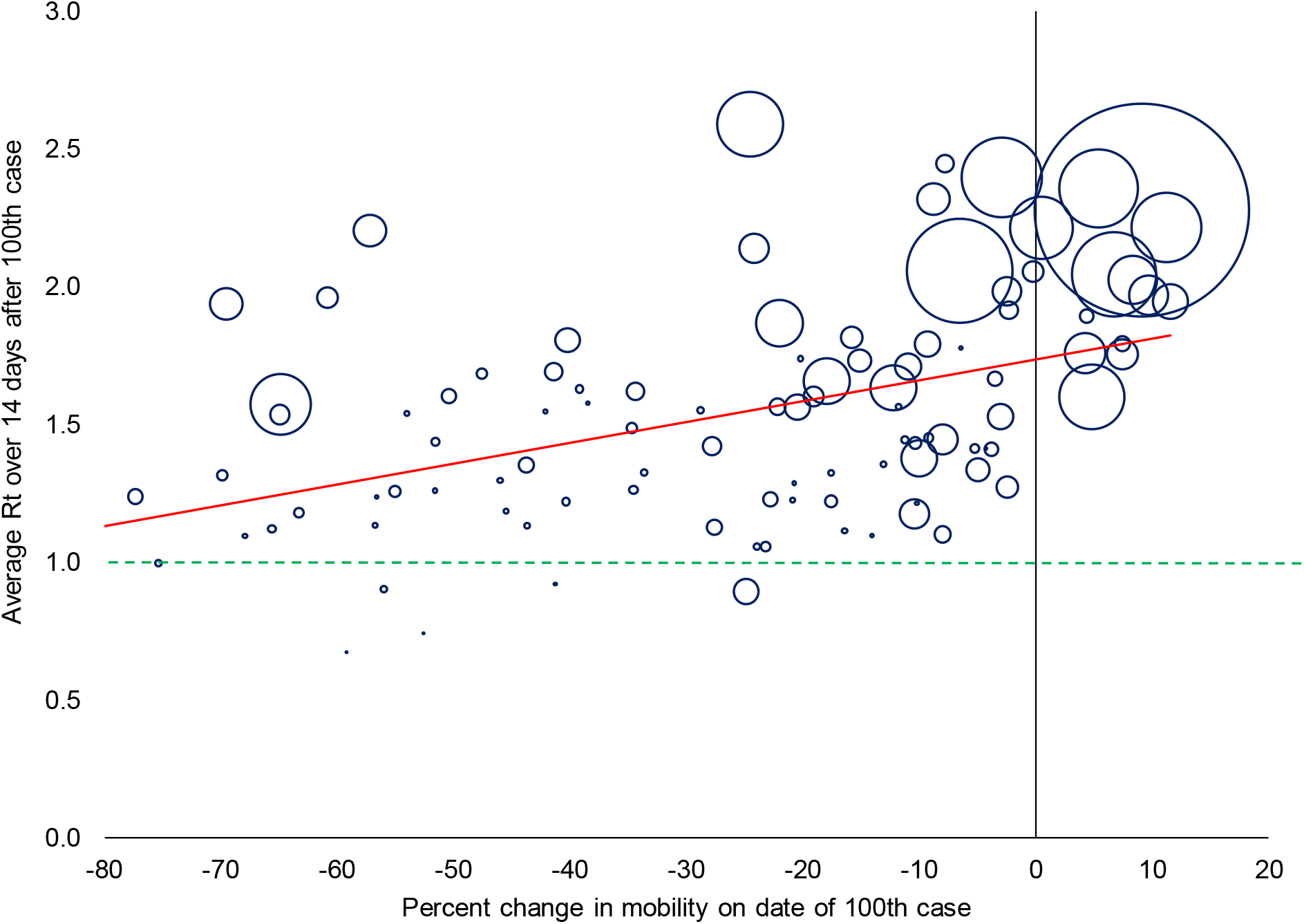
Percent change in mobility on the date of the 100^th^ case and average reproduction numbers in the following two weeks. The time-varying reproduction number R_t_ is the expected number of secondary cases generated by a primary case at time t. The change in mobility (average of retail and recreation, grocery and pharmacy, parks, transit and workplaces) is relative to the baseline median value of the corresponding day of the week during January 3 to February 6, 2020; data for 103 countries from Google Community Mobility Trends Reports. Each bubble represents a country, and the size of the bubble is proportional to the total number of reported cases as of May 28, 2020. The red solid line is the best linear fit of the relationship between the stringency level on the date of the 100^th^ reported case and the average R_t_ in the following two weeks. The green dashed line is the R_t_ threshold: a value below one indicates that a sustained outbreak is unlikely if the measures remain in place.

**Table S1.** Physical distancing measures.

| Physical distancing measure | No. of countries with measures | Average date of implementation | Average days after 1<sup>st</sup> case | Average days after 100<sup>th</sup> case |
| --- | --- | --- | --- | --- |
| Restrictions on international travel | 167 | February 22, 2020 | -10.8 | -33.4 |
| Cancel public events | 165 | March 11, 2020 | 8.3 | -14.6 |
| School closures | 167 | March 12, 2020 | 9.3 | -13.5 |
| Restrictions on size of gatherings | 157 | March 17, 2020 | 13.8 | -9.1 |
| Workplace closures | 155 | March 17, 2020 | 14.9 | -7.6 |
| Restrictions on internal movement | 155 | March 19, 2020 | 16.6 | -6.2 |
| Stay-at-home orders | 152 | March 22, 2020 | 18.5 | -5.0 |
| Public transport closures | 130 | March 22, 2020 | 18.5 | -4.0 |

Author's calculations based on the Oxford COVID-19 Government Response Tracker (OxCGRT). Data of 170 countries as of May 28, 2020.

**Table S2.**
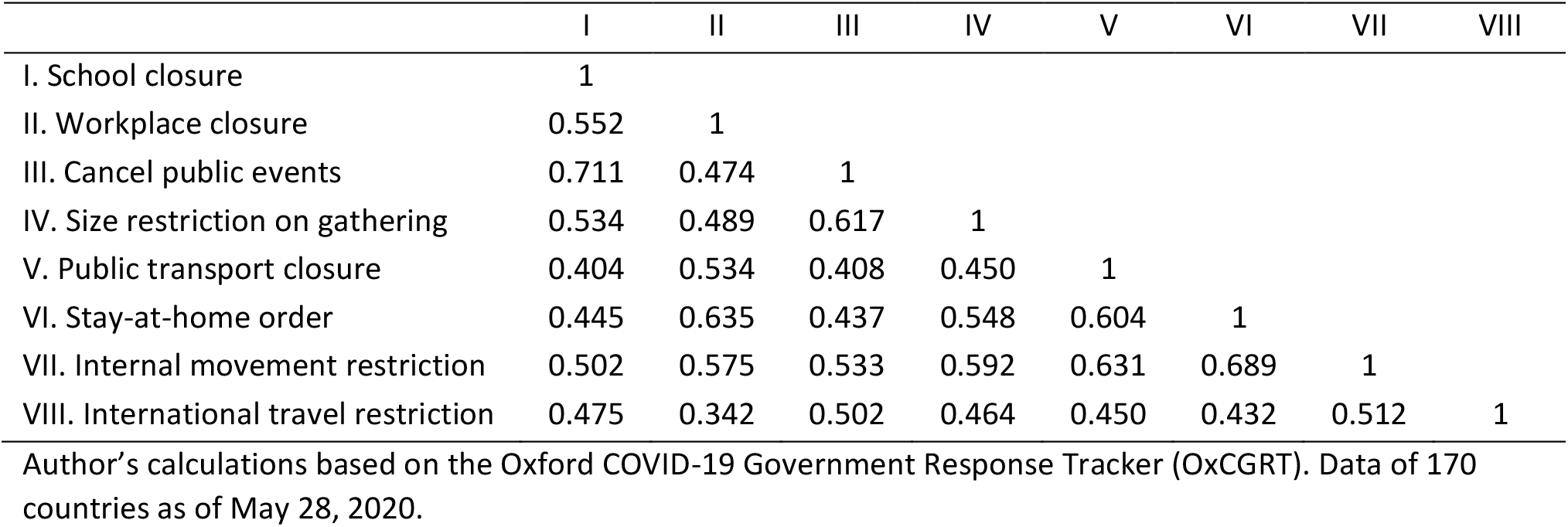
Pairwise correlation of physical distancing measures.

|  | I | II | III | IV | V | VI | VII | VIII |
| --- | --- | --- | --- | --- | --- | --- | --- | --- |
| I. School closure | 1 |  |  |  |  |  |  |  |
| II. Workplace closure | 0.552 | 1 |  |  |  |  |  |  |
| III. Cancel public events | 0.711 | 0.474 | 1 |  |  |  |  |  |
| IV. Size restriction on gathering | 0.534 | 0.489 | 0.617 | 1 |  |  |  |  |
| V. Public transport closure | 0.404 | 0.534 | 0.408 | 0.450 | 1 |  |  |  |
| VI. Stay-at-home order | 0.445 | 0.635 | 0.437 | 0.548 | 0.604 | 1 |  |  |
| VII. Internal movement restriction | 0.502 | 0.575 | 0.533 | 0.592 | 0.631 | 0.689 | 1 |  |
| VIII. International travel restriction | 0.475 | 0.342 | 0.502 | 0.464 | 0.450 | 0.432 | 0.512 | 1 |

Author's calculations based on the Oxford COVID-19 Government Response Tracker (OxCGRT). Data of 170 countries as of May 28, 2020.

**Table S3.** Estimated impact of the type of physical distancing measures on COVID-19 transmission.

|  | (1) | (2) | (3) | (4) |
| --- | --- | --- | --- | --- |
| Restrictions on international travel |  |  |  |  |
| 1 | -0.2287<br>(0.2299) |  |  | -0.3312<br>(0.2209) |
| 2 | -0.4322<br>(0.5014) |  |  | -0.5259<br>(0.4968) |
| 3 | 0.3088<br>(0.2759) |  |  | 0.3184<br>(0.2813) |
| 4 | -0.3192<br>(0.2371) |  |  | -0.2829<br>(0.2319) |
| 5 | -0.1270<br>(0.182) |  |  | -0.0403<br>(0.1795) |
| 6 | -0.2306<br>(0.1636) |  |  | -0.1872<br>(0.1674) |
| 7 | -0.4069**<br>(0.1618) |  |  | -0.2500<br>(0.1731) |
| 8 | -0.5393***<br>(0.1691) |  |  | -0.3815**<br>(0.1775) |
| Restrictions on mass gatherings |  |  |  |  |
| 1 |  | -0.1673<br>(0.2002) |  | -0.0474<br>(0.2043) |
| 2 |  | N/A |  | N/A |
| 3 |  | -0.0738<br>(0.1477) |  | 0.1158<br>(0.1810) |
| 4 |  | -0.3960**<br>(0.1621) |  | -0.0997<br>(0.1902) |
| 5 |  | -0.1712<br>(0.1906) |  | 0.0713<br>(0.2198) |
| 6 |  | -0.5214***<br>(0.1497) |  | -0.1476<br>(0.2027) |
| Lockdown |  |  |  |  |
| 1 |  |  | 0.0769<br>(0.1522) | -0.1082<br>(0.1742) |
| 2 |  |  | -0.5917**<br>(0.2432) | -0.8266***<br>(0.2590) |
| 3 |  |  | -0.1475<br>(0.1805) | -0.2515<br>(0.1918) |
| 4 |  |  | -0.5442**<br>(0.2190) | -0.4082*<br>(0.2363) |
| 5 |  |  | -0.1845<br>(0.1328) | -0.1395<br>(0.1671) |
| 6 |  |  | -0.4961***<br>(0.1166) | -0.3690**<br>(0.1591) |
| In GDP per capita | -0.0231<br>(0.0396) | -0.0411<br>(0.041) | -0.0477<br>(0.0404) | -0.0647<br>(0.0424) |
| In population density | -0.0225<br>(0.0315) | -0.0207<br>(0.0308) | -0.0091<br>(0.0296) | -0.0089<br>(0.0311) |
| % age 65 in population | -0.0028<br>(0.0105) | 0.0033<br>(0.0102) | 0.0062<br>(0.0100) | 0.0030<br>(0.0101) |
| Temperature | -0.0210***<br>(0.0059) | -0.0197***<br>(0.0054) | -0.0173***<br>(0.0053) | -0.0170***<br>(0.0057) |
| Constant | 2.6568***<br>(0.3766) | 2.7066***<br>(0.3933) | 2.6327***<br>(0.3739) | 3.0237***<br>(0.4297) |
| Number of countries | 142 | 142 | 142 | 142 |
| R<sup>2</sup> | 0.334 | 0.334 | 0.375 | 0.470 |
| Adjusted R<sup>2</sup> | 0.272 | 0.289 | 0.327 | 0.366 |

**Table S4.** Estimated impact of the type of physical distancing measures on growth of COVID-19 cases.

|  | (1) | (2) | (3) | (4) |
| --- | --- | --- | --- | --- |
| Restrictions on international travel |  |  |  |  |
| 1 | -11.5241*<br>(6.4055) |  |  | -13.9037**<br>(6.3857) |
| 2 | -4.3420<br>(13.9714) |  |  | -6.8805<br>(14.3637) |
| 3 | -3.1768<br>(7.6889) |  |  | -1.5921<br>(8.1343) |
| 4 | -11.5567*<br>(6.6056) |  |  | -9.7012<br>(6.7039) |
| 5 | -6.7669<br>(5.0718) |  |  | -4.3670<br>(5.1914) |
| 6 | -8.9413*<br>(4.5579) |  |  | -6.2495<br>(4.8393) |
| 7 | -14.9550**<br>(4.5095) |  |  | -9.4285*<br>(5.0047) |
| 8 | -15.3598**<br>(4.7113) |  |  | -9.8495*<br>(5.1321) |
| Restrictions on mass gatherings |  |  |  |  |
| 1 |  | -11.3489**<br>(5.5239) |  | -8.0236<br>(5.9083) |
| 2 |  | N/A |  | N/A |
| 3 |  | -6.4666<br>(4.0747) |  | -0.1768<br>(5.2319) |
| 4 |  | -14.0559***<br>(4.4727) |  | -6.2775<br>(5.4983) |
| 5 |  | -10.5430**<br>(5.2583) |  | -2.8170<br>(6.3564) |
| 6 |  | -15.9865***<br>(4.1313) |  | -5.9179<br>(5.8615) |
| Lockdown |  |  |  |  |
| 1 |  |  | -3.0011<br>(4.2961) | -6.9561<br>(5.0370) |
| 2 |  |  | -10.9708<br>(6.8614) | -14.9819**<br>(7.4892) |
| 3 |  |  | 0.2952<br>(5.0943) | -2.7972<br>(5.5463) |
| 4 |  |  | -16.5760***<br>(6.1800) | -11.5052*<br>(6.8333) |
| 5 |  |  | -8.4072**<br>(3.7467) | -7.0555<br>(4.8320) |
| 6 |  |  | -12.7560***<br>(3.2912) | -10.0471**<br>(4.5988) |
| In GDP per capita | 0.2485<br>(1.1024) | -0.0591<br>(1.1325) | -0.4026<br>(1.1398) | -0.8913<br>(1.2256) |
| In population density | 0.0542<br>(0.8772) | 0.2200<br>(0.8492) | 0.1598<br>(0.8366) | 0.3009<br>(0.9004) |
| % age 65 in population | -0.0689<br>(0.2924) | 0.0031<br>(0.2811) | 0.0784<br>(0.2826) | 0.0468<br>(0.2917) |
| Temperature | -0.4738***<br>(0.1651) | -0.4930***<br>(0.1483) | -0.4551***<br>(0.1503) | -0.3903**<br>(0.1643) |
| Constant | 27.4590***<br>(10.4931) | 29.1527**<br>(10.8533) | 27.9928**<br>(10.5507) | 41.3448***<br>(12.4237) |
| Number of countries | 142 | 142 | 142 | 142 |
| R<sup>2</sup> | 0.310 | 0.323 | 0.335 | 0.408 |
| Adjusted R<sup>2</sup> | 0.245 | 0.277 | 0.284 | 0.293 |

**Table S5.** Predicted time-varying reproduction numbers on date of the 100^th^ case.

| Country | SI | TR | MG | LD | Predicted $R_t$ | Estimated $R_t$<br>using<br>reported data | Difference | Total cases as<br>of May 28,<br>2020 |
| --- | --- | --- | --- | --- | --- | --- | --- | --- |
| Madagascar | 95.37 | 7 | 4 | 6 | 1.34 | 0.74 | -0.60 | 612 |
| Japan | 25.00 | 6 | 1 | 0 | 1.87 | 1.27 | -0.59 | 16,651 |
| Myanmar | 86.11 | 6 | 6 | 6 | 1.31 | 0.74 | -0.56 | 206 |
| Finland | 42.59 | 6 | 3 | 1 | 1.93 | 1.41 | -0.52 | 6,692 |
| Brunei | 52.78 | 8 | 4 | 0 | 1.25 | 0.73 | -0.52 | 141 |
| Ethiopia | 73.15 | 4 | 6 | 6 | 1.30 | 0.83 | -0.47 | 731 |
| Egypt | 18.52 | 0 | 0 | 3 | 1.80 | 1.33 | -0.47 | 19,666 |
| Iceland | 16.67 | 4 | 0 | 0 | 1.98 | 1.53 | -0.45 | 1,805 |
| Kuwait | 74.07 | 7 | 5 | 3 | 1.33 | 0.89 | -0.44 | 23,267 |
| Thailand | 33.80 | 3 | 3 | 0 | 1.89 | 1.45 | -0.44 | 3,054 |
| Vietnam | 50.00 | 8 | 3 | 5 | 1.51 | 1.10 | -0.41 | 327 |
| Guam | 67.59 | 5 | 6 | 6 | 1.09 | 0.68 | -0.41 | 171 |
| Greece | 40.74 | 0 | 3 | 3 | 1.82 | 1.41 | -0.40 | 2,903 |
| Sweden | 0.00 | 0 | 0 | 0 | 2.15 | 1.75 | -0.40 | 35,088 |
| Bahrain | 25.00 | 1 | 0 | 0 | 1.47 | 1.10 | -0.37 | 9,633 |
| Eswatini | 82.41 | 8 | 4 | 5 | 1.40 | 1.04 | -0.36 | 272 |
| Slovenia | 28.70 | 5 | 4 | 1 | 1.68 | 1.32 | -0.36 | 1,471 |
| Liberia | 79.63 | 6 | 6 | 6 | 1.33 | 0.97 | -0.36 | 266 |
| Iraq | 77.78 | 5 | 6 | 5 | 1.58 | 1.22 | -0.36 | 5,135 |
| Aruba | 82.41 | 6 | 6 | 6 | 1.09 | 0.74 | -0.35 | 101 |
| Norway | 11.11 | 0 | 0 | 0 | 2.13 | 1.79 | -0.34 | 8,383 |
| Rwanda | 100.00 | 8 | 6 | 6 | 1.25 | 0.92 | -0.33 | 346 |
| Senegal | 66.67 | 7 | 6 | 5 | 1.37 | 1.06 | -0.32 | 3,253 |
| Estonia | 44.44 | 1 | 3 | 5 | 1.76 | 1.44 | -0.32 | 1,840 |
| Jordan | 100.00 | 7 | 5 | 6 | 1.39 | 1.09 | -0.30 | 720 |
| Trinidad and Tobago | 79.63 | 8 | 6 | 6 | 0.96 | 0.67 | -0.29 | 116 |
| Andorra | 31.48 | 0 | 1 | 6 | 1.51 | 1.25 | -0.26 | 763 |
| Kosovo | 92.59 | 7 | 6 | 6 | 1.42 | 1.17 | -0.25 | 1,047 |
| Bermuda | 96.30 | 8 | 6 | 6 | 0.95 | 0.71 | -0.25 | 139 |
| Algeria | 36.11 | 5 | 3 | 0 | 1.86 | 1.62 | -0.24 | 8,857 |
| Tanzania | 46.30 | 8 | 4 | 0 | 1.52 | 1.29 | -0.24 | 509 |
| Taiwan | 27.78 | 8 | 1 | 0 | 1.45 | 1.21 | -0.24 | 441 |
| Bulgaria | 70.37 | 6 | 6 | 6 | 1.49 | 1.26 | -0.23 | 2,460 |
| Russia | 60.65 | 6 | 4 | 5 | 2.01 | 1.82 | -0.19 | 370,680 |
| Venezuela | 82.41 | 7 | 4 | 6 | 1.09 | 0.90 | -0.18 | 1,245 |
| Cuba | 66.67 | 6 | 3 | 5 | 1.61 | 1.43 | -0.18 | 1,974 |
| Uruguay | 54.63 | 5 | 3 | 2 | 1.35 | 1.18 | -0.16 | 803 |
| Mozambique | 50.93 | 4 | 4 | 1 | 1.57 | 1.41 | -0.16 | 227 |
| Slovak Republic | 75.00 | 7 | 4 | 4 | 1.36 | 1.21 | -0.16 | 1,515 |
| Ghana | 52.78 | 8 | 4 | 0 | 1.36 | 1.23 | -0.13 | 7,303 |
| Burkina Faso | 84.26 | 7 | 3 | 6 | 1.36 | 1.23 | -0.13 | 845 |
| Afghanistan | 62.04 | 1 | 3 | 6 | 1.55 | 1.42 | -0.13 | 12,456 |
| Democratic Republic<br>of Congo | 77.78 | 8 | 4 | 6 | 1.22 | 1.09 | -0.13 | 2,659 |
| Kyrgyz Republic | 92.13 | 8 | 6 | 6 | 1.46 | 1.33 | -0.13 | 1,594 |
| Mauritius | 82.41 | 7 | 3 | 6 | 1.30 | 1.18 | -0.12 | 334 |
| Azerbaijan | 68.52 | 6 | 6 | 5 | 1.67 | 1.56 | -0.12 | 4,568 |
| Hong Kong | 45.37 | 6 | 3 | 2 | 1.23 | 1.11 | -0.11 | 1,066 |
| Albania | 84.26 | 7 | 4 | 6 | 1.43 | 1.32 | -0.10 | 1,050 |
| San Marino | 83.33 | 7 | 6 | 4 | 1.18 | 1.08 | -0.10 | 667 |
| Bosnia and<br>Herzegovina | 89.81 | 5 | 6 | 6 | 1.54 | 1.44 | -0.10 | 2,435 |
| Denmark | 35.19 | 3 | 3 | 1 | 2.01 | 1.92 | -0.10 | 11,480 |
| Tunisia | 87.96 | 5 | 5 | 6 | 1.51 | 1.41 | -0.10 | 1,051 |
| Georgia | 100.00 | 8 | 6 | 6 | 1.35 | 1.26 | -0.09 | 735 |
| Pakistan | 52.78 | 6 | 5 | 0 | 1.85 | 1.76 | -0.09 | 61,227 |
| Niger | 61.11 | 7 | 4 | 5 | 1.45 | 1.36 | -0.09 | 955 |
| Cape Verde | 75.93 | 8 | 3 | 6 | 1.32 | 1.24 | -0.09 | 390 |
| Uzbekistan | 90.74 | 7 | 3 | 6 | 1.60 | 1.52 | -0.09 | 3,333 |
| Lebanon | 52.78 | 8 | 1 | 4 | 1.21 | 1.13 | -0.08 | 1,161 |
| Qatar | 30.56 | 6 | 1 | 0 | 1.44 | 1.38 | -0.07 | 48,947 |
| Guyana | 90.74 | 8 | 6 | 6 | 1.07 | 1.01 | -0.07 | 139 |
| Kazakhstan | 80.09 | 7 | 6 | 5 | 1.62 | 1.56 | -0.06 | 9,576 |
| Indonesia | 43.52 | 6 | 3 | 1 | 1.59 | 1.53 | -0.06 | 23,851 |
| India | 48.15 | 6 | 3 | 1 | 1.65 | 1.60 | -0.05 | 158,333 |
| Argentina | 88.89 | 7 | 5 | 5 | 1.59 | 1.54 | -0.05 | 13,920 |
| Paraguay | 93.52 | 7 | 6 | 6 | 1.18 | 1.13 | -0.05 | 884 |
| Kenya | 83.33 | 8 | 6 | 6 | 1.11 | 1.06 | -0.05 | 1,471 |
| Sri Lanka | 97.22 | 8 | 6 | 6 | 1.03 | 0.99 | -0.04 | 1,469 |
| Philippines | 75.00 | 6 | 3 | 5 | 1.63 | 1.60 | -0.03 | 15,049 |
| Bolivia | 96.30 | 7 | 6 | 6 | 1.27 | 1.24 | -0.03 | 7,768 |
| Canada | 22.22 | 2 | 3 | 0 | 2.02 | 2.02 | 0.00 | 87,508 |
| Costa Rica | 71.30 | 7 | 3 | 6 | 1.29 | 1.30 | 0.01 | 984 |
| Czech Republic | 57.41 | 6 | 3 | 3 | 1.81 | 1.82 | 0.01 | 9,086 |
| United Kingdom | 11.11 | 0 | 0 | 0 | 2.04 | 2.05 | 0.01 | 267,240 |
| Chile | 47.22 | 0 | 3 | 5 | 1.86 | 1.87 | 0.01 | 82,289 |
| Congo | 97.22 | 8 | 4 | 6 | 1.12 | 1.13 | 0.01 | 571 |
| Mexico | 2.78 | 1 | 0 | 0 | 1.62 | 1.63 | 0.02 | 78,023 |
| United Arab Emirates | 45.37 | 6 | 3 | 2 | 1.15 | 1.17 | 0.03 | 31,969 |
| Peru | 94.44 | 7 | 5 | 5 | 1.55 | 1.57 | 0.03 | 135,905 |
| Austria | 19.44 | 5 | 0 | 0 | 2.02 | 2.05 | 0.03 | 16,515 |
| Puerto Rico | 93.52 | 6 | 6 | 6 | 1.15 | 1.18 | 0.03 | 3,397 |
| Poland | 57.41 | 7 | 3 | 3 | 1.76 | 1.81 | 0.04 | 22,473 |
| Cyprus | 92.59 | 7 | 5 | 6 | 1.30 | 1.35 | 0.04 | 939 |
| Honduras | 100.00 | 7 | 6 | 6 | 1.21 | 1.26 | 0.04 | 4,640 |
| El Salvador | 88.89 | 8 | 4 | 6 | 1.07 | 1.12 | 0.05 | 2,109 |
| Mali | 72.22 | 5 | 4 | 6 | 1.27 | 1.32 | 0.06 | 1,116 |
| Somalia | 56.48 | 8 | 4 | 1 | 1.38 | 1.44 | 0.06 | 1,731 |
| Ireland | 48.15 | 0 | 5 | 1 | 1.73 | 1.79 | 0.06 | 24,803 |
| Netherlands | 2.78 | 0 | 0 | 1 | 1.88 | 1.95 | 0.06 | 45,768 |
| Serbia | 81.48 | 8 | 6 | 5 | 1.55 | 1.62 | 0.07 | 11,275 |
| Singapore | 25.00 | 6 | 1 | 0 | 1.38 | 1.45 | 0.07 | 32,876 |
| South Africa | 55.56 | 6 | 3 | 1 | 1.64 | 1.71 | 0.07 | 25,937 |
| Romania | 61.11 | 6 | 4 | 3 | 1.65 | 1.73 | 0.08 | 18,594 |
| Sierra Leone | 80.56 | 8 | 4 | 6 | 1.17 | 1.26 | 0.09 | 782 |
| Guatemala | 93.52 | 6 | 6 | 6 | 1.22 | 1.31 | 0.09 | 4,145 |
| Lithuania | 81.48 | 7 | 6 | 6 | 1.45 | 1.55 | 0.10 | 1,647 |
| Nigeria | 82.87 | 8 | 6 | 6 | 1.02 | 1.13 | 0.10 | 8,733 |
| South Sudan | 92.59 | 8 | 6 | 6 | 1.11 | 1.22 | 0.11 | 806 |
| Malaysia | 22.22 | 6 | 1 | 0 | 1.54 | 1.67 | 0.13 | 7,619 |
| Switzerland | 25.00 | 0 | 4 | 0 | 1.85 | 1.98 | 0.14 | 30,678 |
| Saudi Arabia | 68.52 | 7 | 3 | 5 | 1.51 | 1.66 | 0.14 | 78,541 |
| Central African<br>Republic | 68.52 | 8 | 4 | 6 | 1.21 | 1.36 | 0.15 | 702 |
| Hungary | 67.59 | 7 | 4 | 4 | 1.34 | 1.49 | 0.15 | 3,816 |
| Cote d'Ivoire | 80.56 | 7 | 4 | 6 | 1.18 | 1.33 | 0.15 | 2,556 |
| Moldova | 87.04 | 7 | 6 | 6 | 1.44 | 1.60 | 0.16 | 7,537 |
| Morocco | 90.74 | 7 | 4 | 6 | 1.38 | 1.55 | 0.17 | 7,601 |
| Panama | 51.85 | 6 | 3 | 1 | 1.51 | 1.69 | 0.18 | 11,728 |
| Cameroon | 57.41 | 5 | 4 | 4 | 1.25 | 1.43 | 0.19 | 5,436 |
| Uganda | 87.04 | 6 | 4 | 6 | 1.39 | 1.58 | 0.19 | 281 |
| Gabon | 87.04 | 8 | 6 | 6 | 1.03 | 1.22 | 0.19 | 2,319 |
| New Zealand | 75.93 | 8 | 6 | 5 | 1.36 | 1.56 | 0.20 | 1,154 |
| Belgium | 13.89 | 1 | 0 | 0 | 1.76 | 1.97 | 0.21 | 57,592 |
| Guinea | 73.15 | 7 | 4 | 6 | 1.22 | 1.44 | 0.22 | 3,446 |
| Croatia | 66.67 | 8 | 5 | 6 | 1.40 | 1.63 | 0.23 | 2,244 |
| Nepal | 92.59 | 8 | 6 | 6 | 1.30 | 1.54 | 0.24 | 886 |
| Luxembourg | 79.63 | 0 | 6 | 6 | 1.44 | 1.68 | 0.25 | 4,001 |
| Colombia | 50.93 | 3 | 4 | 2 | 1.31 | 1.56 | 0.25 | 24,104 |
| Australia | 19.44 | 6 | 0 | 0 | 1.63 | 1.89 | 0.26 | 7,139 |
| Belarus | 8.33 | 5 | 0 | 0 | 2.06 | 2.32 | 0.26 | 38,956 |
| Palestine | 94.44 | 7 | 6 | 6 | 1.27 | 1.54 | 0.26 | 613 |
| Brazil | 42.13 | 3 | 3 | 1 | 1.77 | 2.06 | 0.29 | 411,821 |
| Ukraine | 88.89 | 7 | 6 | 6 | 1.48 | 1.77 | 0.29 | 21,584 |
| Zambia | 65.28 | 8 | 6 | 5 | 1.44 | 1.74 | 0.30 | 1,057 |
| France | 19.44 | 4 | 3 | 0 | 1.87 | 2.21 | 0.34 | 145,746 |
| Ecuador | 93.52 | 7 | 3 | 5 | 1.59 | 1.94 | 0.34 | 38,103 |
| Chad | 88.89 | 8 | 4 | 6 | 1.09 | 1.45 | 0.36 | 715 |
| Benin | 59.72 | 4 | 6 | 5 | 1.41 | 1.78 | 0.37 | 339 |
| Portugal | 32.41 | 5 | 0 | 3 | 1.77 | 2.14 | 0.37 | 31,292 |
| Jamaica | 78.70 | 4 | 6 | 6 | 1.18 | 1.55 | 0.37 | 569 |
| South Korea | 45.37 | 5 | 3 | 0 | 2.07 | 2.45 | 0.38 | 11,344 |
| Oman | 81.48 | 8 | 6 | 6 | 0.97 | 1.35 | 0.38 | 8,373 |
| Spain | 11.11 | 0 | 0 | 0 | 2.02 | 2.40 | 0.38 | 236,769 |
| Dominican Republic | 81.48 | 7 | 3 | 5 | 1.55 | 1.96 | 0.41 | 15,723 |
| Israel | 52.78 | 8 | 4 | 0 | 1.39 | 1.82 | 0.42 | 16,793 |
| China | 8.33 | 0 | 0 | 0 | 2.32 | 2.75 | 0.43 | 84,106 |
| United States | 11.11 | 5 | 1 | 0 | 1.83 | 2.28 | 0.45 | 1,699,933 |
| Sudan | 81.48 | 8 | 4 | 6 | 1.09 | 1.57 | 0.48 | 4,146 |
| Djibouti | 94.44 | 7 | 6 | 6 | 1.15 | 1.66 | 0.51 | 2,697 |
| Iran | 18.52 | 0 | 3 | 1 | 1.87 | 2.40 | 0.54 | 141,591 |
| Germany | 25.00 | 1 | 1 | 0 | 1.62 | 2.21 | 0.59 | 179,717 |
| Italy | 69.91 | 6 | 5 | 5 | 1.74 | 2.36 | 0.62 | 231,139 |
| Turkey | 58.33 | 6 | 3 | 3 | 1.78 | 2.59 | 0.81 | 158,762 |
| Bangladesh | 87.04 | 6 | 6 | 6 | 1.25 | 2.20 | 0.96 | 38,292 |

